# Impaired language in Alzheimer’s disease: A comparison between English and Persian implicates content-word frequency rather than the noun-verb distinction

**DOI:** 10.1101/2024.04.09.24305534

**Authors:** Mahya Sanati, Sabereh Bayat, Mehrdad Mohammad Panahi, Amirhossein Khodadadi, Sahar Rezaee, Mahdieh Ghasimi, Sara Besharat, Zahra Mahboubi Fooladi, Mostafa Almasi Dooghaee, Morteza Sanei Taheri, Bradford C Dickerson, Adele Goldberg, Neguine Rezaii

## Abstract

This study challenges the conventional psycholinguistic view that the distinction between nouns and verbs is pivotal in understanding language impairments in neurological disorders. Traditional views link frontal brain region damage with verb processing deficits and posterior temporoparietal damage with noun difficulties. However, this perspective is contested by findings from patients with Alzheimer’s disease (pwAD), who show impairments in both word classes despite their typical temporoparietal atrophy. Notably, pwAD tend to use semantically lighter verbs in their speech than healthy individuals. By examining English-speaking pwAD and comparing them with Persian-speaking pwAD, this research aims to demonstrate that language impairments in Alzheimer’s disease (AD) stem from the distributional properties of words within a language rather than distinct neural processing networks for nouns and verbs. We propose that the primary deficit in AD language production is an overreliance on high-frequency words. English has a set of particularly high-frequency verbs that surpass most nouns in usage frequency. Since pwAD tend to use high-frequency words, the byproduct of this word distribution in the English language would be an over-usage of high-frequency verbs. In contrast, Persian features complex verbs with an overall distribution lacking extremely high-frequency verbs like those found in English. As a result, we hypothesize that Persian-speaking pwAD would not have a bias toward the overuse of high-frequency verbs.

We analyzed language samples from 95 English-speaking pwAD and 91 healthy controls, along with 27 Persian-speaking pwAD and 27 healthy controls. Employing uniform automated natural language processing methods, we measured the usage rates of nouns, verbs, and word frequencies across both cohorts.

Our findings showed that English-speaking pwAD use higher-frequency verbs than healthy individuals, a pattern not mirrored by Persian-speaking pwAD. Crucially, we found a significant interaction between the frequencies of verbs used by English and Persian speakers with and without AD. Moreover, regression models that treated noun and verb frequencies as separate predictors did not outperform models that considered overall word frequency alone in classifying AD.

In conclusion, this study suggests that language abnormalities among English-speaking pwAD reflect the unique distributional properties of words in English rather than a universal noun-verb class distinction. Beyond offering a new understanding of language abnormalities in AD, the study highlights the critical need for further investigation across diverse languages to deepen our insight into the mechanisms of language impairments in neurological disorders.

## Introduction

The distinction between nouns and verbs is considered central to language.^1,2^ While a few languages may lack distinct classes of adjectives, prepositions, or adverbs,^3–5^ it is unclear that any language entirely abolishes the distinction between nouns and verbs. A noun-verb distinction may be inevitable for functional reasons. Effective communication typically requires a subject to talk about (usually denoted by nouns) and some comment about that subject (often conveyed through verbs)^1^. Owing to its centrality, many language impairments have been examined through the lens of the noun and verb distinction. This approach has led to the conventional perspective that patients with frontal lobe lesions produce fewer verbs, while those with posterior brain lesions produce fewer nouns. This pattern of impairment is often attributed to the assumed neurobiological functions of these word classes, with verbs processed in areas associated with actions near the motor cortex,^6,7^ and nouns linked to the posterior sensory association areas.^8–16^ However, more detailed psycholinguistic analysis of the language produced by people with brain damage indicates this conventional understanding may be oversimplified. For instance, although patients with frontal lobe damage typically use fewer verbs, the verbs they produce are often semantically richer or “heavier” than unimpaired controls.^17,18^

Another complex pattern is evident among people with Alzheimer’s disease (pwAD), who typically have predominant neurodegeneration in temporoparietal regions. The conventional view predicts pwAD should show greater deficits with nouns than verbs. However, pwAD have deficits in production and comprehension of both nouns and verbs compared to healthy individuals, and they display worse performance on verbs relative to nouns at the single-word level.^19–22^ In connected discourse, pwAD tend to rely on nonspecific and semantically light verbs during sentence production.^14,23^ In explaining why pwAD over-rely on nonspecific verbs, Kim et al. proposed that the deficit stems from the semantic complexity associated with heavier verbs.^23^ In particular, more specific verbs, such as “walk” or “ski”, may well be associated with more specific nouns, such as “shoes” or “boots,” compared to less specific verbs, such as “go”. As the specific noun associates degrade in pwAD, verb retrieval becomes more challenging. Therefore, according to this view, access to verbs is indirectly affected by the loss of specific nouns rather than through a direct impairment in verb access.

In the current work, we test a different hypothesis: the language spoken by pwAD is not directly influenced by the distinction between nouns and verbs but is instead a byproduct of content word frequency in a particular language. A condition such as AD, which makes lexical access more difficult, is recognized to involve an overreliance on high-frequency words compared to controls, who are better able to access words that specifically suit their intended messages.^24–26^ We propose that English-speaking AD patients over-rely on certain verbs, an observation replicated here, because of the distributional properties of verbs in English. In particular, English has a set of very high-frequency main verbs (e.g., *be, do, have, know, go, get, think, come, see, take)*, which are more frequent than nearly any noun (e.g., *people, time*, or *thing*) (Figure 1). English also has a large set of low-frequency verbs, partly due to the existence of Latinate synonyms for common verb meanings (e.g., *exist, act, acquire, understand, possess, consider, arrive, visualize*) and partly because English verbs often encode manner (e.g., *waltz, shuffle, saunter)* or a resultant state *(redden, flatten, amuse*). Therefore, examining verb impairment only in English makes it challenging to disentangle various contributing factors—such as word class and word frequency—that may underlie the observed language features in AD. In this context, an investigation of AD speech in a language with different distributional properties than English is used to distinguish the role of grammatical category (nouns vs. verbs) and frequency.

**Figure 1.**
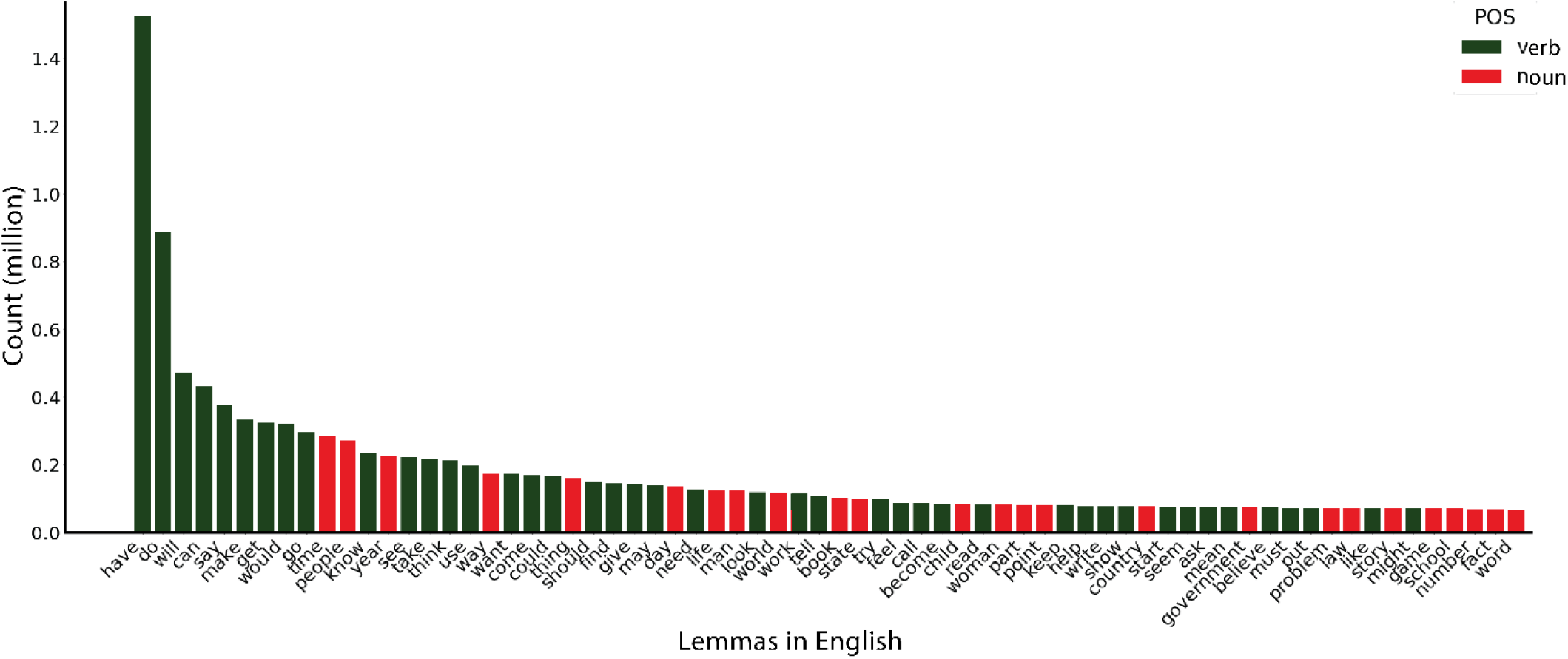
Distribution of verb and noun lemmas in English based on the spoken modality of The Corpus of Contemporary American English (COCA)^27^. Most of the highest frequency words are semantically light verbs.

For these reasons, we conducted this study to compare the production of verbs and nouns by English speakers with that of Persian speakers. Like English, Persian (also known as Farsi) has light verbs that can appear as main verbs, but Persian’s light verbs are commonly used as part of complex verbs. Complex verbs consist of a non-verb element, the host, followed by a light verb.^28–30^ The nature of Persian verbs results in a wide variety of complex verbs instead of a collection of high-frequency main verbs, as in English. Thus, a comparative analysis of Persian and English provides a unique vantage point that allows us to ask whether English-speaking pwAD may over-rely on certain verbs because they are particularly frequent rather than because of a difference between nouns and verbs. If the noun-verb distinction is an explanatory factor in language impairment among pwAD, we would expect consistent challenges with verbs (or nouns) to be evident across both languages. We hypothesized that the data would not support this possibility. Instead, the core problem in pwAD is access to low-frequency, semantically richer words, which would impact lexical classes differently in languages with different distributional properties of word classes.

## Methods

### Participants

#### The English cohort

We obtained English samples from DementiaBank, a component of the TalkBank project.^31^ The dataset was collected at the University of Pittsburgh as part of the Alzheimer Research Program between 1983 and 1988 during a 5-year-follow up (comprehensive information available in Becker et al. 1994).^32^ Inclusion criteria consisted of being older than 44 years old, having at least seven years of education, not having previous neurologic disorders, not taking neuroleptic drugs, having at least a score of 10 in Mini-Mental State Exam (MMSE) and being able to give informed consent. Participants were assessed using comprehensive examinations, including neuropsychiatric batteries, laboratory data, CT scans, and EEGs. From this dataset, we included 95 patients with probable AD and 91 age-matched healthy individuals and included each person’s first language sample if more than one was collected. We selected the subset of pwAD from the DementiaBank in such a way that they match the MMSE and age of pwAD in the Persian cohort as closely as possible (Table 1).

**Table 1.**
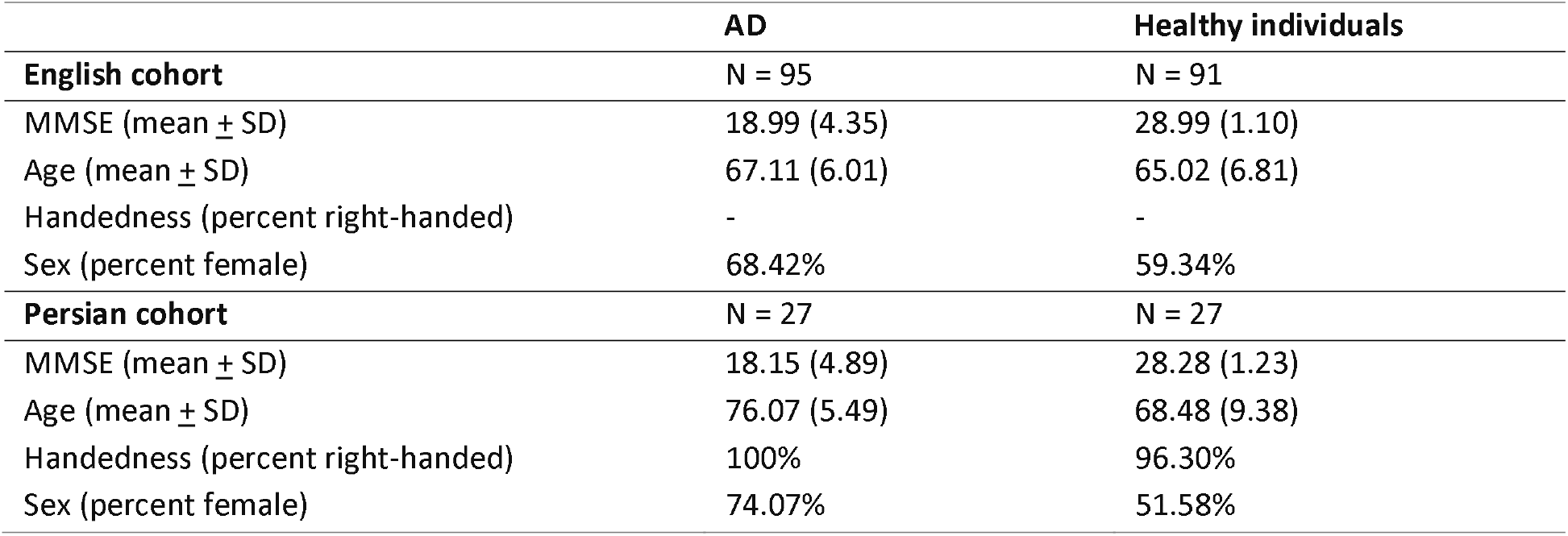
Demographic and clinical characteristics of pwAD and healthy speakers of English and Persian.

|  | **AD** | **Healthy individuals** |
| --- | --- | --- |
| **English cohort** | **N = 95** | **N = 91** |
| MMSE (mean ± SD) | 18.99 (4.35) | 28.99 (1.10) |
| Age (mean ± SD) | 67.11 (6.01) | 65.02 (6.81) |
| Handedness (percent right-handed) | - | - |
| Sex (percent female) | 68.42% | 59.34% |
| **Persian cohort** | **N = 27** | **N = 27** |
| MMSE (mean ± SD) | 18.15 (4.89) | 28.28 (1.23) |
| Age (mean ± SD) | 76.07 (5.49) | 68.48 (9.38) |
| Handedness (percent right-handed) | 100% | 96.30% |
| Sex (percent female) | 74.07% | 51.58% |

#### The Persian cohort

Twenty-seven Persian-speaking pwAD and 27 age-matched healthy individuals were recruited from the Brain and Cognition Clinic in Tehran, Iran. A complete clinical history was taken from patients and their caregivers. Demographic features, clinical presentations, medical, and family histories were included in the interviews. A complete clinical examination was performed, emphasizing the assessment of motor features such as parkinsonism, praxis, language and speech, gait, and balance. The cognitive examination included Addenbrook’s Cognitive Examination-Persian version (ACE)^33^ and Mini-Mental Status Exam (MMSE).^34^ Blood tests and the brain MRI of all patients were reviewed to confirm the diagnosis and rule out other medical conditions. The blood tests included complete blood count, biochemistry, renal and liver function tests, vitamin B12, 25-OH-D3, thyroid function test, syphilis, and HIV serologic test. Mild or Major Neurocognitive Disorders and Alzheimer’s disease were diagnosed by DSM-5 and NINCDS-ADRDA criteria^35^, respectively. In addition to obtaining a clinical history for all healthy individuals, the neurotypicality was confirmed using MMSE, brain MRI, or both. The research section of the Persian cohort was approved by the ethics committee of Iran University of Medical Sciences, which governs human subjects research in accordance with their guidelines. All participants provided written informed consent to participate in this study.

#### Language samples

Connected speech samples were obtained for both languages by asking participants to describe the Cookie Theft picture, a component of the Boston Diagnostic Aphasia Examination.^36^ The recorded samples were transcribed into text for further analysis.

#### Language features

We used Stanza, an open-source Python natural language processing toolkit that supports 66 human languages, including English and Persian, to extract part-of-speech (POS) tags such as verbs and nouns.^37^ The dependency relation “compound:lvc” in Stanza denotes complex verbs in Persian.

#### Word frequency

Frequencies of both nouns and verbs were calculated for each language on a corpus of word counts extracted from Wikipedia, comprising 4 million words in each language. We opted for Wikipedia for its cross-linguistic consistency. The process of extracting Wikipedia pages adhered to an algorithm that encompassed similar major topics across both languages. The algorithm initiated the extraction of text from eleven distinct topics shared by both languages and extended to include all text accessible through hyperlinks on the respective pages until it reached the desired word count. We used the lemmatized form of words to determine word frequency, excluding the verb “be” and auxiliary verbs from our analysis to be conservative since the effects described below would be even more pronounced if these verbs had been included. Log frequencies are used in the models described below.

## Results

### Analysis of the Wikipedia corpus in English and Persian

The distributions of verbs and nouns in the 4 million word samples in each language are shown in Figure 2. The highest-frequency verbs in English are higher frequency than those in Persian and far higher than other verbs in either language. For instance, the cumulative count of the top 20 verbs in English (shaded area) reaches 1,187,341, whereas in Persian, the top 20 verbs accumulate a total of 697,890 counts. On the other hand, the distribution of nouns in the two languages is more similar. The cumulative count of the top 20 nouns is 2,702,463 in English and 3,196,274 in Persian.

**Figure 2.**
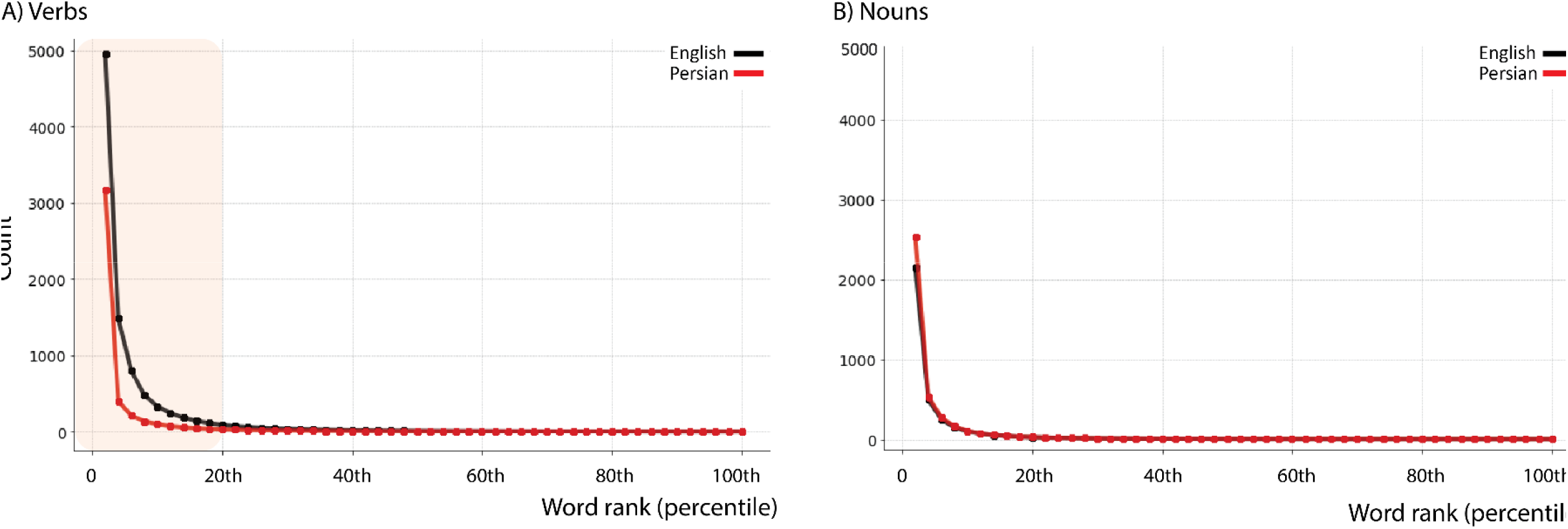
The distributions of verbs (A) and nouns (B) in English and Persian.

**Figure 3.**
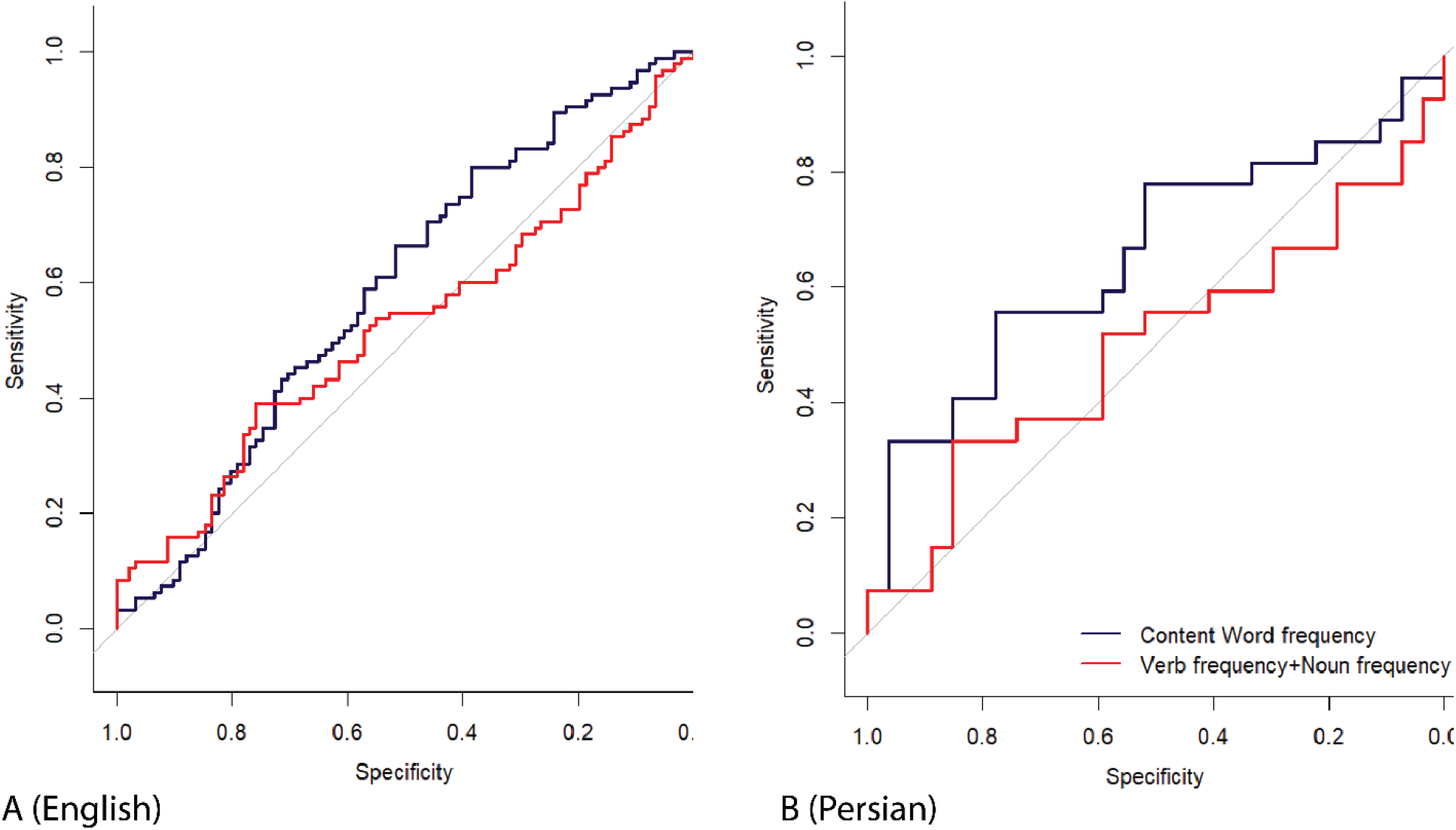
A comparison of ROC curves for predicting AD based on word frequency versus distinguishing the frequency of nouns and verbs in English (A) and Persian (B) shows no advantage for the separation of word classes.

### Analysis of language samples of pwAD and healthy controls across English and Persian

First, we measure the noun-verb ratios in both languages to test for a disproportionate deficit in one or the other word class among pwAD compared to healthy individuals. We used the following formula to obtain the noun-verb ratio in each language sample: total number of nouns/(total number of nouns + total number of verbs).

Results show no significant difference in the ratio of noun-verb use between pwAD and healthy individuals in either language: English (pwAD, *M* = 0.54; healthy *M* = 0.57; *t* = 1.53, *p* = 0.128); Persian (pwAD, *M* = 0.70; healthy *M* = 0.71; *t* = 0.324, *p* = 0.75).

There was a significant interaction between the frequencies of verbs used by English and Persian speakers with and without AD (*ß*= -0.638, *p* = 0.015), so we compared speakers with and without AD in each language. Consistent with prior literature^19^, English-speaking pwAD favored higher (log) frequency verbs (Mean = 7.03) in comparison to healthy individuals (M = 6.76), (*t* =-2.595, *p* = 0.010). Critically, the log frequency of verbs used by Persian-speaking pwAD (M = 7.22) was *not* higher than verbs used by healthy Persian speakers (*M* = 7.60) (*t* = 1.108, *p* = 0.27).

We also compared the frequencies of nouns by English and Persian speakers with and without AD. Here we find an interaction showing the opposite pattern: Persian speakers’ production of nouns was more influenced by AD than English speakers’ production of nouns (ß= 0.448, *p* = 0.009). However, direct comparisons between noun frequencies used by pwAD and healthy individuals did not reach significance in either language: In English speakers: pwAD (*M* = 5.13) and healthy individuals (*M* =5.26) *t* = 1.715, *p* = 0.088; in Persian speakers pwAD (6.19) and healthy individuals (5.88) (t = -1.925, p = 0.060).

We next sought to evaluate whether the distinction of verb frequency and noun frequency as two separate variables offers an advantage in classifying AD compared to content word frequency as a single variable. For this, we generated two models. The first model consisted of verb frequency and noun frequency as two separate predictor variables to classify AD, while the second model consisted of content word frequency as a single predictor variable. We analyzed the ROC curves for each model, and the AUC was used as a measure of discriminative ability. Results indicated no evidence to claim that the distinction between nouns and verbs offers an advantage over content word frequency in predicting AD. Specifically, we used the DeLong test to statistically compare two AUCs from correlated ROC curves. In the English cohort, the DeLong test yielded a Z-score of 1.312 (p = 0.190). The AUC for content word frequency was 0.589, and the AUC for the two-variable model, which consisted of verb frequency and noun frequency, was 0.524. Similarly, in the Persian cohort, the DeLong test yielded a Z-score of 1.876 (p = 0.060). The AUC for content word frequency was 0.653, marginally higher than the AUC for the two-variable model consisting of verb frequency and noun frequency, which was 0.498.

## Discussion

In this study, we explored language production in pwAD to address discrepancies between emerging findings on noun and verb processing in this population and the longstanding views on the brain’s segregated processing networks for the two word classes. According to the conventional view, grammatical class information is crucial in language production and comprehension.^38–40^ The formulation of these views has evolved and consolidated in tandem with observations from patients with various forms of aphasia. Particularly noteworthy were findings viewed as a double dissociation between word class and aphasia type. Aphasic patients with temporoparietal damage were found to exhibit more pronounced difficulties in producing nouns than verbs, while patients with Broca’s aphasia with lesions in left frontal regions showed the opposite pattern.^41–43^ These observations have been construed as compelling evidence for the claim that lexical knowledge in the brain is organized by grammatical class.^44^ This conceptualization posits a division within neural networks, with left temporal regions dedicated to noun representation and left frontal areas specialized for verb representation.^45,46^

However, this view has proven overly simplistic as it fails to account for several emerging observations from patients with various neurological deficits. For example, while nonfluent aphasic patients with left frontal lobe damage exhibit a lower ratio of verbs to nouns in language production, they often produce semantically complex or heavy verbs.^17,18^ The verbs that nonfluent patients produce can be even heavier than those produced by healthy individuals, indicating that a simple verb class cannot provide a comprehensive account of the linguistic characteristics observed in the disease.^47–49^ Another notable discrepancy is language production in pwAD. The traditional understanding of word processing across separate neural networks predicts that pwAD would struggle more with noun production than verb production, given the predominance of posterior temporal and parietal neurodegeneration and the relative sparing of frontal areas, particularly in the early stages of the disease. Reflecting the prevailing notions, early studies primarily focused on nouns in this patient population.^50^ However, accumulating findings challenge this prediction, with some outcomes being completely contrary, such as the one we focus on here. Specifically, English-speaking pwAD use verbs of higher frequencies than controls,^51^ and this frequency effect in verb naming has been shown to worsen as the disease progresses.^52^

In this work, we adopt a cross-linguistic approach to test an alternative hypothesis about language changes in pwAD, which is that the fundamental difficulty is in accessing less frequent (therefore more specific and informative) words of any lexical class. This hypothesis posits that word class effects are simply a reflection of the distributional properties of words in a given language. Our investigation suggests that the well-documented overreliance on high-frequency verbs in English-speaking pwAD compared to healthy controls stems from their particularly skewed distribution in English. English features a group of especially high-frequency verbs, such as “take” and “get,” which may emerge as a distinct subset within the broader category of verbs upon cluster analyses based on frequency.^53^ These high-frequency verbs possess abstract and flexible meanings as illustrated by phrases like “take a walk,” “take a break,” and “take a bath,” which convey very distinct actions.^54,55^ Conversely, in Persian, light verbs are more commonly integrated into complex verb structures rather than combined with independent noun phrases as in the English examples just mentioned. As predicted, if frequency distinguishes verbs and nouns more strongly in English than in Persian, the observation of increased use of high-frequency verbs by English-speaking pwAD did not hold for Persian-speaking pwAD, whose preference for higher-frequency words did not disproportionately affect verbs. Our analysis confirmed a significant interaction between verb frequencies among English and Persian speakers with and without AD. The interaction undermines the notion of segregated processing for nouns and verbs as the explanatory factor and instead indicates that English-speaking patients’ overreliance on high-frequency verbs stems from English verbs’ distributional properties. We further showed that a regression model that incorporates verb frequency and noun frequency as two distinct predictors offers no advantage over a model with content word frequency as a single predictor in classifying people as AD or controls.

What emerges as a common finding across both languages is the reliance of pwAD on higher-frequency words, with differing consequences dependent on the distributional properties of the language spoken. Basic principles of information theory confirm that higher-frequency words tend to offer less information due to their higher predictability within context.^56^ This reliance on high-frequency words seems to be invariant to word class, with word class effects being a byproduct of how nouns and verbs are distributed within a given language. It is well-established that the language of individuals with AD often exhibits vagueness,^57–60^, attributed to the use of uninformative high-frequency words.^19,61^ The direction of causation still remains unclear: Either individuals with AD struggle to access low-frequency words, resulting in uninformative language, or they experience vague thoughts that hinder the selection of specific and informative words.

The new insight into language abnormalities of AD – that implicates word frequency rather than word class distinction-- is consistent with our recent theoretical framework about nonfluent aphasia, known as the Information Restoration Paradigm.^62^ This framework also argued that language impairments could be explained in terms of frequency, except that nonfluent aphasiac patients favor lower-frequency words over higher-frequency words.^47,48,62–64^ We analyzed the language of nonfluent patients—with both Broca’s and the nonfluent variant of primary progressive aphasia— who, consistent with existing literature, produce higher proportions of content to function words, heavy verbs to light verbs, and nouns to verbs. Rather than interpreting these findings as isolated symptoms, we identified a common thread: low frequency. Across all three cases of word class dissociation, we observed that patients with nonfluent aphasia consistently opted for the lower-frequency word type. We interpreted this pattern as reflecting the patients’ economic behavior in language production. Confronted with challenges in generating long and complex sentences, patients gravitated towards using low-frequency words as a compensatory strategy since low-frequency words inherently offer more informational value than high-frequency words. Consequently, these patients tend to avoid high-frequency verbs and rely on nouns, along with low-frequency verbs, due to their informational advantage. This pattern of language production in nonfluent aphasia presents a mirror image of the phenomenon observed in AD, where patients struggle with producing informative messages.

Therefore, our interpretation, grounded in word frequency rather than word class distinction, offers a theoretical framework capable of reconciling the two incongruent findings present in the literature on the pattern of word use in AD and nonfluent aphasia. This interpretation is consistent with views that consider nouns and verbs to be processed through the same shared neural networks.^65,66^ These views, initially introduced by Sapir (1921)^1^ and later refined through functionalist approaches to language processing,^67^ suggest that grammatical class emerges as a consequence of distribution and semantics.^66^ Even small recurrent networks can predict the grammatical properties of words and categorize them based on their grammatical class in a probabilistic way on the basis of their distribution.^68^ In this framework, lexical categories include prototypical and less prototypical exemplars which self-organize based on distribution, semantics, and pragmatics.^66,69–71^

Future studies are needed to investigate diverse languages with varying distributional properties within their lexicons to better understand the root psycholinguistic causes of impairments in pwAD. For instance, unlike findings in which English-speaking pwAD exhibited better performance in single-word production of nouns compared to verbs, a study of Chinese speakers with AD showed similar performance across both nouns and verbs.^72^ Adopting cross-linguistic approaches is crucial to avoid presumptions about certain linguistic features being language universals or biologically determined when, in reality, they may be contingent upon the properties of specific languages. The comparative methodology further enhances our understanding of the nuances in diverse languages and, critically, allows for new insights into the causes of impaired language in different groups of people.

## Availability of data and materials

The English dataset was obtained from the DementiaBank repository. The Persian dataset obtained for this study, in addition to the computational codes in Python, can be accessed upon contacting the senior author at

## Competing interests

The authors declare no competing interests to declare.

## Funding

This work was supported by Alzheimer’s Association Clinician Scientist Fellowship (AACSF) 2022A015154 and MGH Screening Technologies in Primary Care Innovation Fund (PCIF) 2023A063002, as well as National Institutes of Health grants R21 DC019567, R21 AG073744, and R01 NS131395.

## Authors’ contributions

SB, MS, MMP, AK, MG, SR, SB, ZM, and MST participated in the data collection. SB, MS, MMP, AK, MG, SR, and MAD contributed to patients’ clinical and neuropsychological assessment. NR and AG conceptualized the idea and were major contributors to writing the manuscript. NR and AG analyzed data. NR, AG, and BCD edited the manuscript. All authors contributed to the initial writing and read and approved the final manuscript.

## Acknowledgment

We thank Harvard Catalyst, The Harvard Clinical and Translational Science Center (National Center for Research Resources and the National Center for Advancing Translational Sciences, National Institutes of Health Award UL1 TR002541) for its biostatistician consultation service. We also thank Mahan Rezaei for transcriptional services in Persian.

## Notes

### Competing Interest Statement

The authors have declared no competing interest.

### Author Declarations

The research section of the Persian cohort was approved by the ethics committee of Iran University of Medical Sciences, which governs human subjects research in accordance with their guidelines. All participants provided written informed consent to participate in this study.

## References

1. Sapir, E. Language an Introduction to the Study of Speech. (Harcourt, Brace, New York, 1921).

2. Meillet, A. Linguistique Historique et Linguistique Generale. (Honore Champion, 1926).

3. Roots of Human Sociality: Culture, Cognition and Interaction. (Routledge, New York, NY, 2006).

4. Hengeveld, K. Parts of Speech. in Layered Structure and Reference in a Functional Perspective: Papers from the Functional Grammar Conference, Copenhagen, 1990 (eds. Fortescue, M., Harder, P. & Kristoffersen, L.) 29 (John Benjamins Publishing Company, 1992). doi:10.1075/pbns.23.04hen.

5. Croft, W. Radical Construction Grammar: Syntactic Theory in Typological Perspective. (Oxford University Press, Oxford⍰; New York, 2001).

6. Ash, S. et al. Nonfluent Speech in Frontotemporal Lobar Degeneration. J. Neurolinguistics 22, 370–383 (2009).

7. Grossman, M. & Irwin, D. J. Primary Progressive Aphasia and Stroke Aphasia. Contin. Minneap. Minn 24, 745–767 (2018).

8. Marcotte, K. et al. Verb production in the nonfluent and semantic variants of primary progressive aphasia: the influence of lexical and semantic factors. Cogn. Neuropsychol. 31, 565–583 (2014).

9. Wilson, S. M. et al. Connected speech production in three variants of primary progressive aphasia. Brain Lond. Engl. s1878 133, 2069–2088 (2010).

10. Auclair-Ouellet, N., Fossard, M., Macoir, J. & Laforce, R. The Nonverbal Processing of Actions Is an Area of Relative Strength in the Semantic Variant of Primary Progressive Aphasia. J. Speech Lang. Hear. Res. JSLHR 63, 569–584 (2020).

11. Breedin, S. D., Saffran, E. M. & Coslett, H. B. Reversal of the concreteness effect in a patient with semantic dementia. Cogn. Neuropsychol. 11, 617–660 (1994).

12. Migliaccio, R. et al. The brain network of naming: A lesson from primary progressive aphasia. PLoS ONE 11, (2016).

13. Bates, E., Chen, S., Tzeng, O. J., Li, P. & Opie, M. The noun-verb problem in Chinese aphasia. Brain Lang. 41, 203–233 (1991).

14. Bates, E., Harris, C., Marchman, V., Wulfeck, B. & Kritchevsky, M. Production of Complex Syntax in Normal Ageing and Alzheimer’s Disease. Lang. Cogn. Process. - LANG Cogn. PROCESS 10, 487–539 (1995).

15. Petersen, S. E., Fox, P. T., Posner, M. I., Mintun, M. & Raichle, M. E. Positron emission tomographic studies of the cortical anatomy of single-word processing. Nature 331, 585–589 (1988).

16. Martin, A., Wiggs, C. L., Ungerleider, L. G. & Haxby, J. V. Neural correlates of category-specific knowledge. Nature 379, 649–652 (1996).

17. Gordon, J. K. & Dell, G. S. Learning to divide the labor: an account of deficits in light and heavy verb production. Cogn. Sci. 27, 1–40 (2003).

18. Breedin, S. D., Saffran, E. M. & Schwartz, M. F. Semantic Factors in Verb Retrieval: An Effect of Complexity. Brain Lang. 63, 1–31 (1998).

19. Williams, E., McAuliffe, M. & Theys, C. Language changes in Alzheimer’s disease: A systematic review of verb processing. Brain Lang. 223, 105041 (2021).

20. Cotelli, M. et al. Action and object naming in frontotemporal dementia, progressive supranuclear palsy, and corticobasal degeneration. Neuropsychology 20, 558–565 (2006).

21. Druks, J. et al. Is action naming better preserved (than object naming) in Alzheimer’s disease and why should we ask? Brain Lang. 98, 332–340 (2006).

22. Almor, A. et al. A Common Mechanism in Verb and Noun Naming Deficits in Alzheimer’s Patients. Brain Lang. 111, 8–19 (2009).

23. Kim, M. & Thompson, C. K. Verb deficits in Alzheimer’s disease and agrammatism: Implications for lexical organization. Brain Lang. 88, 1–20 (2004).

24. Gayraud, F.Lee, H.-R. & Barkat-Defradas, M. Syntactic and lexical context of pauses and hesitations in the discourse of Alzheimer patients and healthy elderly subjects. Clin. Linguist. Phon. 25, 198–209 (2011).

25. Bridges, K. A. & Van Lancker Sidtis, D. Formulaic language in Alzheimer’s disease. Aphasiology 27, 799–810 (2013).

26. Kavé, G. & Goral, M. Word retrieval in connected speech in Alzheimer’s disease: a review with meta-analyses. Aphasiology 32, 1–23 (2017).

27. Davies, M. The Corpus of Contemporary American English (COCA). (2008).

28. Goldberg, A. E. Words by default: The Persian complex predicate construction. Mismatch Form-Funct. Incongruity Archit. Gramm. 1, 17–146 (2003).

29. Karimi-Doostan, G. Light verbs and Structural case.

30. Sharif, B. The Evolution of Complex Predicates in Iranian Languages. Lang. Sci. 10, 365–400 (2024).

31. MacWhinney, B. The Talkbank Project. in Creating and Digitizing Language Corpora: Volume 1: Synchronic Databases (eds. Beal, J. C., Corrigan, K. P. & Moisl, H. L. 163–180 (Palgrave Macmillan UK, London, 2007). doi:10.1057/9780230223936_7.

32. Becker, J. T., Boller, F., Lopez, O. L., Saxton, J. & McGonigle, K. L. The natural history of Alzheimer’s disease. Description of study cohort and accuracy of diagnosis. Arch. Neurol. 51, 585–594 (1994).

33. Pouretemad, H. R., Khatibi, A., Ganjavi, A., Shams, J. & Zarei, M. Validation of Addenbrooke’s cognitive examination (ACE) in a Persian-speaking population. Dement. Geriatr. Cogn. Disord. 28, 343–347 (2009).

34. Ansari, N. N., Naghdi, S., Hasson, S., Valizadeh, L. & Jalaie, S. Validation of a Mini-Mental State Examination (MMSE) for the Persian population: a pilot study. Appl. Neuropsychol. 17, 190–195 (2010).

35. McKhann, G. et al. Clinical diagnosis of Alzheimer’s disease. Neurology 34, 939–939 (1984).

36. Goodglass, H. & Kaplan, E. Boston Diagnostic Aphasia Examination (BDAE). in (Lea and Febiger. Distributed by Psychological Assessment Resources, Odessa, FL, 1983).

37. Qi, P., Zhang, Y., Zhang, Y., Bolton, J. & Manning, C. D. Stanza: A Python Natural Language Processing Toolkit for Many Human Languages. in Proceedings of the 58th Annual Meeting of the Association for Computational Linguistics: System Demonstrations 101–108 (Association for Computational Linguistics, Online, 2020). doi:10.18653/v1/2020.acl-demos.14.

38. Kempen, G. & Hoenkamp, E. An Incremental Procedural Grammar for Sentence Formulation. Cogn. Sci. 11, 201–258 (1987).

39. Pickering, M. J. & Branigan, H. P. The Representation of Verbs: Evidence from Syntactic Priming in Language Production. J. Mem. Lang. 39, 633–651 (1998).

40. Levelt, W. J. M. Speaking: From Intention to Articulation. (A Bradford Book, Cambridge, MA, USA, 1989).

41. Miceli, G., Silveri, M. C., Villa, G. & Caramazza, A. On the basis for the agrammatic’s difficulty in producing main verbs. Cortex J. Devoted Study Nerv. Syst. Behav. 20, 207–220 (1984).

42. Miceli, G., Silveri, M. C., Nocentini, U. & Caramazza, A. Patterns of dissociation in comprehension and production of nouns and verbs. Aphasiology 2, 351–358 (1988).

43. Zingeser, L. B. & Berndt, R. S. Grammatical class and context effects in a case of pre anomia: implications for models of language processing. Cogn. Neuropsychol 473–516 (1988).

44. Caramazza, A. & Hillis, A. E. Lexical organization of nouns and verbs in the brain. Nature 349, 788–790 (1991).

45. Damasio, A. R. & Tranel, D. Nouns and verbs are retrieved with differently distributed neural systems. Proc. Natl. Acad. Sci. U. S. A. 90, 4957–4960 (1993).

46. Daniele, A., Giustolisi, L., Silveri, M. C., Colosimo, C. & Gainotti, G. Evidence for a possible neuroanatomical basis for lexical processing of nouns and verbs. Neuropsychologia 32, 1325–1341 (1994).

47. Rezaii, N., Ren, B., Quimby, M., Hochberg, D. & Dickerson, B. C. Less is more in language production: an information-theoretic analysis of agrammatism in primary progressive aphasia. Brain Commun. 5, fcad136 (2023).

48. Rezaii, N. et al. Measuring Sentence Information via Surprisal: Theoretical and Clinical Implications in Nonfluent Aphasia. Ann. Neurol. 94, 647–657 (2023).

49. Rezaii, N. An information-theoretic analysis of agrammatism in Broca’s aphasia. 2023.04.23.23288999 Preprint at 10.1101/2023.04.23.23288999 (2023).

50. Cappa, S. F. & Perani, D. The neural correlates of noun and verb processing. J. Neurolinguistics 16, 183–189 (2003).

51. Williams, E., Theys, C. & McAuliffe, M. Lexical-semantic properties of verbs and nouns used in conversation by people with Alzheimer’s disease. PLOS ONE 18, e0288556 (2023).

52. Beber, B. C., da Cruz, A. N. & Chaves, M. L. A behavioral study of the nature of verb production deficits in Alzheimer’s disease. Brain Lang. 149, 128–134 (2015).

53. Rezaii, N. et al. Using Generative Artificial Intelligence to Classify Primary Progressive Aphasia from Connected Speech. 2023.12.22.23300470 Preprint at 10.1101/2023.12.22.23300470 (2023).

54. Jespersen, O. Modern English Grammar on Historical Principles: Part V Syntax. (Allen & Unwin, London, 1965).

55. Kegl, J. Levels of representation and units of access relevant to agrammatism. Brain Lang. 50, 151–200 (1995).

56. Shannon, C. E. A Mathematical Theory of Communication. 55 (1948).

57. Nicholas, M., Obler, L. K., Albert, M. L. & Helm-Estabrooks, N. Empty Speech in Alzheimer’s Disease and Fluent Aphasia. J. Speech Lang. Hear. Res. 28, 405–410 (1985).

58. Obler, L. & Albert, M. Language and aging: A neurobehavioral analysis. Aging Commun. Process. Disord. 107–121 (1981).

59. Snowdon, D. A. et al. Linguistic Ability in Early Life and Cognitive Function and Alzheimer’s Disease in Late Life: Findings From the Nun Study. JAMA 275, 528–532 (1996).

60. Ripich, D. N. & Terrell, B. Y. Patterns of Discourse Cohesion and Coherence in Alzheimer’s Disease. J. Speech Hear. Disord. 53, 8–15 (1988).

61. Ahmed, S., de Jager, C. A.Haigh, A.-M. & Garrard, P. Semantic processing in connected speech at a uniformly early stage of autopsy-confirmed Alzheimer’s disease. Neuropsychology 27, 79–85 (2013).

62. Rezaii, N. & Dickerson, B. C. Artificial Intelligence Enables a Paradigm Shift in Understanding Nonfluent Aphasia. (2024) doi:10.31234/osf.io/wn7vc.

63. Rezaii, N., Ren, B., Quimby, M., Hochberg, D. & Dickerson, B. Less is more in language production: Shorter sentences contain more informative words. 2022.06.02.22275938 Preprint at 10.1101/2022.06.02.22275938 (2022).

64. Josephy-Hernandez, S. et al. Automated analysis of written language in the three variants of primary progressive aphasia. Brain Commun. 5, fcad202 (2023).

65. Crepaldi, D., Berlingeri, M., Paulesu, E. & Luzzatti, C. A place for nouns and a place for verbs? A critical review of neurocognitive data on grammatical-class effects. Brain Lang. 116, 33–49 (2011).

66. Vigliocco, G., Vinson, D. P., Druks, J., Barber, H. & Cappa, S. F. Nouns and verbs in the brain: a review of behavioural, electrophysiological, neuropsychological and imaging studies. Neurosci. Biobehav. Rev. 35, 407–426 (2011).

67. Bates, E. & Macwhinney, B. Functionalist approaches to grammar. Child Lang. State Art (1982).

68. Elman, J. L. Finding structure in time. Cogn. Sci. 14, 179–211 (1990).

69. Givon, T. English Grammar. z.engram2 (John Benjamins Publishing Company).

70. Langacker, R. W. Foundations of Cognitive Grammar: Volume I: Theoretical Prerequisites. (Stanford University Press, Stanford, 1987).

71. Croft, W. Syntactic Categories and Grammatical Relations: The Cognitive Organization of Information. (University of Chicago Press, Chicago, IL, 1991).

72. Lai, Y.-H. & Lin, Y.-T. Factors in action-object semantic disorder for Chinese-speaking persons with or without Alzheimer’s disease. J. Neurolinguistics 26, 298–311 (2013).

